# Clinical, cerebrospinal fluid and neuroimaging findings in COVID-19 encephalopathy: a case series

**DOI:** 10.1101/2020.08.28.20181883

**Authors:** Raphael Tuma, Bruno Guedes, Rafael Carra, Bruno Iepsen, Júlia Rodrigues, Antonio Edvan Camelo-Filho, Gabriel Kubota, Maíra Ferrari, Adalberto Studart Neto, Mariana Hiromi Manoel Oku, Sara Terrim, Cesar Castello Branco Lopes, Carlos E. B. Passos Neto, Matheus D. Fiorentino, Julia C. C. Souza, José Pedro S. Baima, Tomás Silva, Iago Perissinotti, Maria da Graça M. Martin, Marcia Gonçalves, Ida Fortini, Jerusa Smid, Tarso Adoni, Leandro Lucato, Ricardo Nitrini, Helio Gomes, Luiz H Castro

## Abstract

**Objective:** To describe the clinical, neurological, neuroimaging and cerebrospinal fluid (CSF) findings associated with encephalopathy in patients admitted to a COVID-19 tertiary reference center.

**Methods:** We retrospectively reviewed records of consecutive patients with COVID-19 evaluated by a consulting neurology team from March 30, 2020 through May 15, 2020.

**Results:** Fifty-five patients with confirmed SARS-CoV-2 were included, 43 of whom showed encephalopathy, and were further divided into mild, moderate and severe encephalopathy groups. Nineteen patients (44%) had undergone mechanical ventilation and received intravenous sedatives. Eleven (26%) patients were on dialysis. Laboratory markers of COVID-19 severity were very common in encephalopathy patients, but did not correlate with the severity of encephalopathy. Thirty-nine patients underwent neuroimaging studies, which showed mostly non-specific changes. One patient showed lesions possibly related to CNS demyelination. Four had suffered an acute stroke. SARS-CoV-2 was detected by RT-PCR in only one of 21 CSF samples. Two CSF samples showed elevated white blood cell count and all were negative for oligoclonal bands. In our case series the severity of encephalopathy correlated with higher probability of death during hospitalization (OR = 5.5 for each increment in the degree of encephalopathy, from absent (0) to mild (1), moderate (2) or severe (3), p<0.001).

**Conclusion:** In our consecutive series with 43 encephalopathy cases, neuroimaging and CSF analysis did not support the role of direct viral CNS invasion or CNS inflammation as the cause of encephalopathy.

## Introduction

COVID-19 is a multisystem disease that usually targets the respiratory and cardiovascular systems. Neurological manifestations are common, occurring in up to 36.4% of cases^1^. The precise mechanism of neurologic involvement in COVID-19 remains incompletely understood, and may include neuroinvasion by severe acute respiratory syndrome coronavirus type 2 (SARS-CoV-2), cytokine storm, hypoxic and vascular injury, as well as endothelial dysfunction^2^. Central nervous system invasion has not been consistently demonstrated.

Encephalopathy is a major complication of SARS-CoV-2 infection^3^. In previous studies, decreased consciousness was observed in 7.5-19.6% of COVID-19 patients^1,4,5^, and in 14.8-38.9% of severe cases^1,5^. Agitation and confusion occur commonly in patients with acute respiratory distress syndrome in COVID-19^6^. In a previous series of consecutive COVID-19 patients evaluated by neurologists in a dedicated COVID-19 tertiary referral hospital, encephalopathy was the leading reason for a neurology consultation^7^.

## Objective

To describe the underlying conditions associated with encephalopathy in patients admitted to a COVID-19 reference center, including systemic illness, intravenous sedative use, cerebrospinal fluid (CSF) and neuroimaging findings.

## Methods

In March 2020, part of the Hospital das Clínicas da FMUSP was designated as a 900 bed (250 ICU) referral center dedicated to clinically suspected or confirmed COVID-19 patients. A neurology consult team consisting of seven neurologists and eight neurology residents (four PGY-2 and four PGY-3) provided on demand evaluations in the ICUs and wards. We retrospectively reviewed records of consecutive patients with COVID-19 evaluated by the neurology team from March 30, 2020 through May 15, 2020.

We selected cases with confirmed COVID-19, by positive SARS-CoV2 RT-PCR in nasopharyngeal swab or tracheal secretions or positive IgM or IgG serology by immunochromatography.

We classified patients in four groups according to the severity of the encephalopathy:

- No encephalopathy – fully awake, preserved sustained and basic attention, without neuropsychiatric disturbances or psychomotor slowing.
- Mild encephalopathy – Awake or easily arousable patient, with preserved basic attention but impaired sustained attention. Patients with preserved sustained attention and level of consciousness, but presenting psychiatric, behavioral symptoms or psychomotor slowing were also included in this group.
- Moderate encephalopathy – Awake or easily arousable patient, with impaired basic and sustained attention.
- Severe encephalopathy – Comatose patients or patients requiring vigorous stimuli to be aroused. Patients with severe psychomotor agitation were also included in this group.

Basic attention was defined as the ability to concentrate on a simple task, such as counting from 1 to 20, reciting the months in direct order or forward-order digit span. Sustained or complex attention described the patient’s ability to focus on a complex task that requires information processing, such as counting from 20 to 1, reciting the months of the year backwards, or reverse-order digit span.

We did not include patients with: pre-existing encephalopathy without significant worsening during the acute illness, encephalopathy occurring immediately after cardiopulmonary resuscitation, incomplete clinical or neurological evaluation and fully awake patients with severe aphasia precluding attention evaluation. Patients still receiving intravenous sedatives were not excluded, since clinicians noted signs that warranted a neurology consultation despite sedation.

### Data collection

We performed a systematic chart review for all enrolled patients collecting clinical data and neurological exam findings.

Outcome and additional neurological diagnoses established during follow-up were checked for all patients on June 1, 17 days after the evaluation period ended. Only laboratory tests collected within three days of the evaluation were included. CSF analysis included interleukin 6 (IL-6) quantification (Electrochemiluminescence – Cobas® e 411, Roche), presence of oligoclonal bands (isoelectric focusing followed by immunoblotting) and CSF-SARS-CoV-2-RT-PCR (Abbot m2000 system Invitrogen Superscript IV and IDT primers and probes with 40 copies/mL as the detection limit). Chest CT findings were classified as suggestive of viral pneumonia or not, based on radiology reports.

Brain computed tomography (CT), CT angiography, magnetic resonance imaging (MRI) and MRI angiography in encephalopathy patients were systematically reviewed by a trained neuroradiologist for acute changes. MRI was acquired in a 1.5T GE scanner, including T1, T2, FLAIR, DWI and- susceptibility weighted images and, when possible, FLAIR and T1-weighted images post gadolinium injection.

The study was approved by the hospital’s ethics committee and is registered at http://plataformabrasil.saude.gov.br, certification number 33563420.0.0000.0068.

## Statistical analysis

We applied ordinal logistic regression with the degree of encephalopathy as the endpoint. Predictors were selected based on previously described mortality predictors in COVID19^8,9^: age, gender, hypertension, diabetes, lymphocyte count and C-reactive protein. We included two additional predictors to account for a potential contribution of neuroinflammation: CSF white blood cell count and total protein. Intravenous sedation was also included as a predictor. In a secondary analysis we applied logistic regression using encephalopathy degree as a predictor and mortality as an endpoint.

## Results

In the period between March 30 and May 15, 1,720 patients were admitted to the COVID-19 dedicated facility, with 1,263 confirmed SARS-CoV-2 infections. As of June 30, 812 of 1,263 (64.3%) COVID-19 patients had been discharged, 412 (32.6%) had died and 39 (3.1%) were still in the hospital. The neurology team evaluated a total of 66 COVID-19 patients. Eleven patients were excluded. Fifty-five patients were included in the final analysis (Flowchart 1). All patients were also included in a previous study that evaluated reasons for neurology consultation in Hospital das Clínicas^7^. Detailed clinical and ancillary data of patients with encephalopathy were not provided in that report.

**Flowchart 1.**
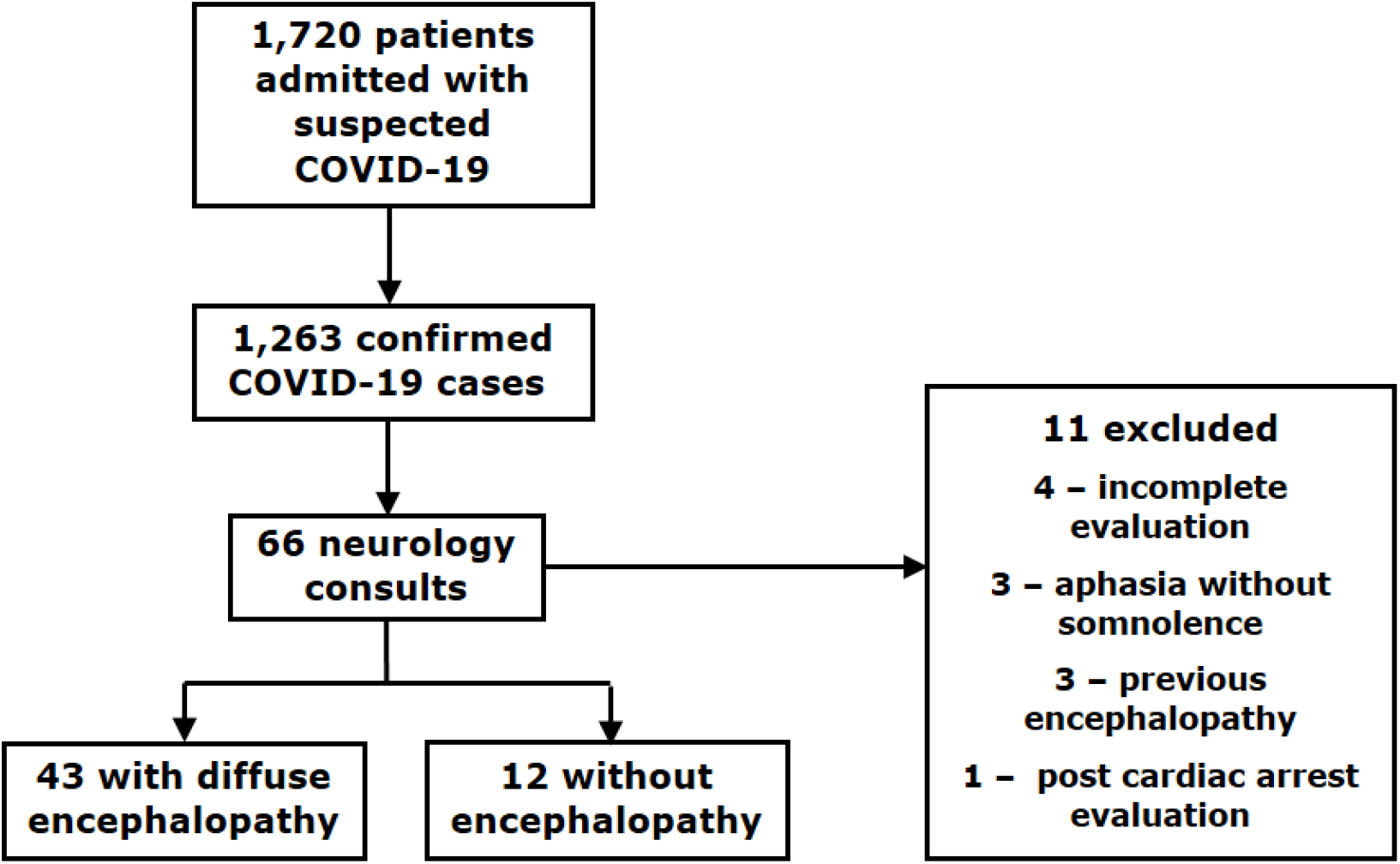
From March 30 to May 15: 1,720 patients admitted to the COVID dedicated facility; 1,263 confirmed SARS-CoV-2 cases throughout the hospital; 66 COVID-19 patients received a neurology consultation, 43 with acute encephalopathy; eleven patients were excluded.

Patients were classified according to encephalopathy severity as: no encephalopathy, 12 cases; mild encephalopathy, 12 cases; moderate encephalopathy, 18 cases; severe encephalopathy, 13 cases.

Demographic data is shown in Table 1, including the median number of days from initial respiratory symptoms to admission and days from admission to neurology consultation. Five patients (12%) developed COVID-19 while already hospitalized for other conditions.

**Table 1.** Demographic data and prior medical history of COVID positive patients with mild, moderate and severe encephalopathy, all encephalopathy cases, and cases without encephalopathy.

|  | **Mild** | **Moderate** | **Severe** | **Encephalopathy Cases** | **Cases without Encephalopathy** |
| --- | --- | --- | --- | --- | --- |
| **Number of cases** | 12 | 18 | 13 | 43 | 12 |
| **Men** | 8 (67%) | 12 (67%) | 9 (69%) | 29 (68%) | 6 (50%) |
| **Age (years)** | 58.5<br>(+/-15.9) | 65.1<br>(+/-12.9) | 58.2<br>(+/-15.8) | 61.2<br>(+/-14.7) | 55.0<br>(+/-9.8) |
| **Over 60 years of age** | 5 (33%) | 11 (61%) | 7 (54%) | 22 (51%) | 3 (25%) |
| **Days from initial respiratory symptoms to admission** | 7.0<br>(5.0 – 12.0) | 2.5<br>(1.3 – 4.8) | 5.0<br>(1.0 – 7.0) | 3.5<br>(2.0 – 7.8) | 6.5<br>(4.5 – 8.0) |
| **Days from admission to initial neurology consultation** | 4.0<br>(1.5 – 6.0) | 10.0<br>(3.3 – 19.8) | 6.0<br>(2.0 – 10.0) | 5.0<br>(3.0 – 16.0) | 11.5<br>(3.3 – 21.8) |
| **Prior medical history** |  |  |  |  |  |
| **Hypertension** | 9 (75%) | 14 (78%) | 8 (62%) | 31 (72%) | 8 (67%) |
| **Diabetes** | 6 (50%) | 8 (44%) | 5 (39%) | 19 (44%) | 4 (33%) |
| **Heart Disease** | 4 (33%) | 2 (11%) | 2 (15%) | 8 (19%) | 0 (0%) |
| **Pulmonary Disease** | 0 (0%) | 2 (11%) | 2 (15%) | 5 (11%) | 1 (8%) |
| **Impaired renal function** | 3 (25%) | 4 (22%) | 4 (31%) | 11 (26%) | 3 (25%) |
| **Alcohol abuse** | 2 (17%) | 1 (6%) | 1 (8%) | 4 (9%) | 0 (0%) |
| **Smoking** | 1 (8%) | 4 (22%) | 5 (39%) | 10 (23%) | 2 (17%) |
| **Cancer (in the previous five years)** | 0 (0%) | 2 (11%) | 3 (23%) | 5 (12%) | 2 (17%) |
| **Immuno-suppressive drug use** | 1 (8%) | 0 (0%) | 3 (23%) | 4 (9%) | 3 (25%) |
| **Prior Neurologic disease** | 6 (50%) | 5 (28%) | 3 (23%) | 14 (33%) | 4 (33%) |
| **Ischemic Stroke** | 4 (33%) | 3 (17%) | 1 (8%) | 8 (19%) | 0 (0%) |
| **Epilepsy** | 3 (25%) | 0 (0%) | 1 (8%) | 4 (9%) | 3 (25%) |
| **Dementia** | 3 (25%) | 1 (6%) | 1 (8%) | 5 (12%) | 0 (0%) |
| Data are median (Interquartile Range), mean (Standard Deviation) or n/N (%). |  |  |  |  |  |

### Clinical Data

Arterial hypertension was the most common comorbidity in the encephalopathy group: 31 of 43 (72%), followed by diabetes in 19 (44%), renal disease in 11 (26%) and smoking in 10 (23%). No patient had HIV or AIDS. Fourteen (33%) had a pre-existing neurologic condition.

Three of 32 (9%) encephalopathy patients reported hyposmia and sixteen of 36 (44%) reported mental status changes, such as somnolence or confusion, prior to admission to the referral center. Three encephalopathy(7%) patients reported seizures before admission, one of whom had a prior history of epilepsy. Three patients in the non-encephalopathy group had a history of epilepsy and presented breakthrough seizures before admission. Of the patients with confirmed COVID-19 and encephalopathy, chest CT was typical of viral infection in 37/39 cases (95%), compared to 8/12 (67%) in the non-encephalopathy group.

Mechanical ventilation, dialysis, vasopressor and sedative use is shown in Table 2. During hospitalization, 19 (44%) patients with encephalopathy received intravenous sedatives for mechanical ventilation, six (14%) of whom were still under sedation on neurological evaluation. All these patients received midazolam and fentanyl. Three of the 19 (16%) also received Propofol, and six (32%) also received Dexmedetomidine. For the 13 patients no longer receiving intravenous sedatives on neurological evaluation, the median number of days from initiation of IV sedatives to IV sedative cessation was 9.0 (IQR 6.0 – 14.0), ranging from three to 42 days. Neurology evaluation occurred a median of 8.0 (IQR 5.0 – 11.0) days after sedation cessation (Table 2).

**Table 2.** Clinical data leading to neurology consultation and patient outcome.

|  | **Mild** | **Moderate** | **Severe** | **Encephalopathy Cases** | **Cases without Encephalopathy** |
| --- | --- | --- | --- | --- | --- |
| **Number of cases** | 12 | 18 | 13 | 43 | 12 |
| **Intravenous sedative use** |  |  |  |  |  |
| **Any time since admission** | 2 (17%) | 10 (56%) | 7 (54%) | 19 (44%) | 4 (33)% |
| **Still in use at time of evaluation** | 0 (0%) | 0 (0%) | 6 (46%) | 6 (14%) | 0 (0%) |
| **Days from sedatives cessation to evaluation** | 4.5<br>(3.8 – 5.3) | 8.0<br>(5.5 – 10.5) | 11.0<br>(11.0–11.0) | 8.0<br>(5.0 – 11.0) | 16.0<br>(11.0 – 18.0) |
| **Days from sedative (Initiation/ withdrawal)** | 11.5<br>(8.8 – 14.3) | 9.5<br>(6.3 – 13.5) | 6.0<br>(6.0 – 6.0) | 9.0<br>(6.0 – 14.0) | 10.0<br>(8.0 – 14.0) |
| **Sedatives in use at evaluation** | 0 (0%) | 0 (0%) | 6 (46%) | 6 (14%) | 0 (0%) |
| **Midazolam** | 2 (100%) | 10 (100%) | 7 (100%) | 19 (100%) | 4 (33%) |
| **Propofol** | 0 (0%) | 1 (10%) | 2 (29%) | 3 (16%) | 0 (0%) |
| **Fentanyl** | 2 (100%) | 10 (100%) | 7 (100%) | 19 (100%) | 4 (33%) |
| **Dexmedetomidine** | 2 (100%) | 1 (10%) | 3 (43%) | 6 (32%) | 1.0 (8%) |
| **Mechanical Ventilation (MV)** |  |  |  |  |  |
| **Any time since admission** | 2 (17%) | 10 (56%) | 7 (54%) | 19 (43%) | 5 (42%) |
| **MV at neurological evaluation** | 0 (0%) | 4 (22%) | 5 (39%) | 9 (21%) | 0 (0%) |
| **Days on MV** | 10.5 (7.75 – 13.25) | 16.0 (13.5 – 19.3) | 13.0 (13.0 – 13.0) | 15.0<br>(13.0 – 17.0) | 13.0<br>(11.0 – 19.0) |
| **Days from MV cessation to initial evaluation** | 5.5<br>(4.75 – 6.25) | 6.5<br>(4.5 – 10.0) | 4.0<br>(4.0 – 4.0) | 6.0<br>(4.0 – 7.0) | 10.0<br>(5.0 – 17.0) |
| **Other clinical data** |  |  |  |  |  |
| **Required IV vasopressors** | 2 (17%) | 8 (44 %) | 8 (62%) | 18 (42%) | 5,0 (42%) |
| **On dialysis** | 2 (17%) | 4 (22%) | 5 (39%) | 11 (26%) | 2,0 (17%) |
| **On antipsychotics** | 4 (33%) | 3 (17%) | 5 (39%) | 12 (28%) | 3 (25%) |
| **Outcome** |  |  |  |  |  |
| **Discharged** | 10 (84%) | 10 (56%) | 1 (8%) | 21 (49%) | 9 (75%) |
| **Death** | 0 (0%) | 5 (28%) | 6 (46%) | 11 (26%) | 1 (8%) |
| Data are median (IQR), mean (SD) or n/N (%). N is the total number of patients with available data and n the number of presenting the finding. |  |  |  |  |  |

Table 3 presents neurological exam findings and clinical data obtained from chart review closest to the neurology consultation. Four (9%) patients had preserved level of consciousness and attention, but were included in the mild encephalopathy group due to marked psychomotor slowing or acute psychiatric symptoms (psychosis and hypervigilance) without previous psychiatric disease.

**Table 3.**
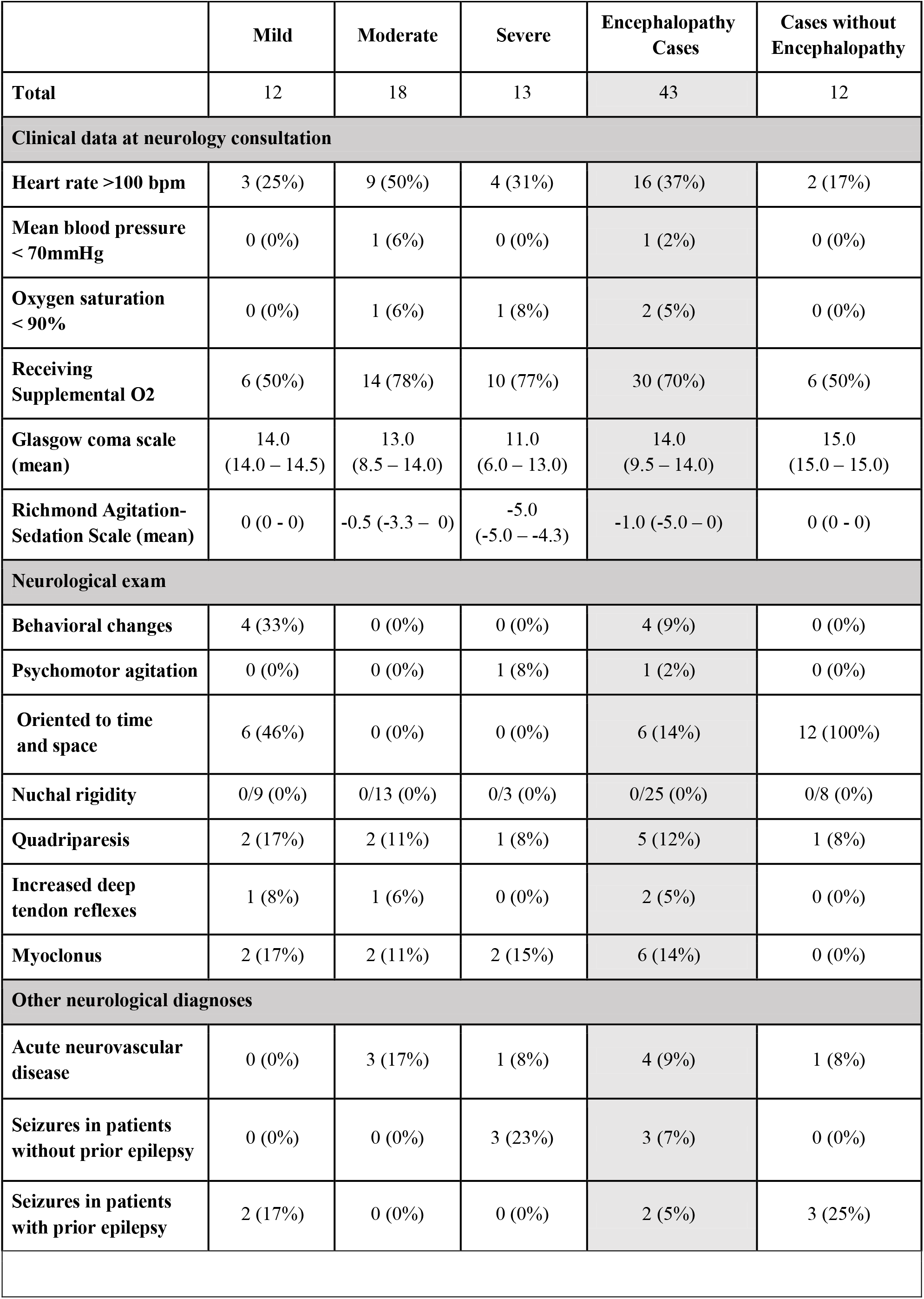
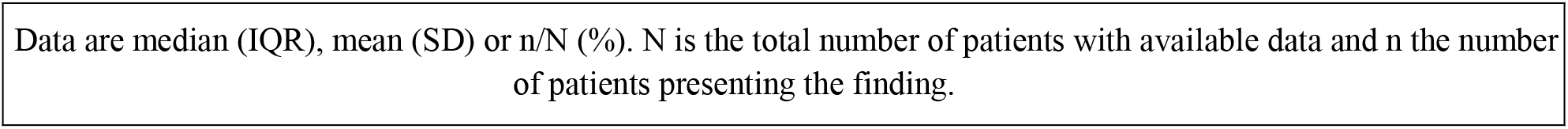
Clinical data from the neurological evaluation.

|  | Mild | Moderate | Severe | Encephalopathy Cases | Cases without Encephalopathy |
| --- | --- | --- | --- | --- | --- |
| **Total** | 12 | 18 | 13 | 43 | 12 |
| **Clinical data at neurology consultation** |  |  |  |  |  |
| **Heart rate >100 bpm** | 3 (25%) | 9 (50%) | 4 (31%) | 16 (37%) | 2 (17%) |
| **Mean blood pressure < 70mmHg** | 0 (0%) | 1 (6%) | 0 (0%) | 1 (2%) | 0 (0%) |
| **Oxygen saturation < 90%** | 0 (0%) | 1 (6%) | 1 (8%) | 2 (5%) | 0 (0%) |
| **Receiving Supplemental O2** | 6 (50%) | 14 (78%) | 10 (77%) | 30 (70%) | 6 (50%) |
| **Glasgow coma scale (mean)** | 14.0<br>(14.0 – 14.5) | 13.0<br>(8.5 – 14.0) | 11.0<br>(6.0 – 13.0) | 14.0<br>(9.5 – 14.0) | 15.0<br>(15.0 – 15.0) |
| **Richmond Agitation-Sedation Scale (mean)** | 0 (0 - 0) | -0.5 (-3.3 – 0) | -5.0<br>(-5.0 – -4.3) | -1.0 (-5.0 – 0) | 0 (0 - 0) |
| **Neurological exam** |  |  |  |  |  |
| **Behavioral changes** | 4 (33%) | 0 (0%) | 0 (0%) | 4 (9%) | 0 (0%) |
| **Psychomotor agitation** | 0 (0%) | 0 (0%) | 1 (8%) | 1 (2%) | 0 (0%) |
| **Oriented to time and space** | 6 (46%) | 0 (0%) | 0 (0%) | 6 (14%) | 12 (100%) |
| **Nuchal rigidity** | 0/9 (0%) | 0/13 (0%) | 0/3 (0%) | 0/25 (0%) | 0/8 (0%) |
| **Quadriparesis** | 2 (17%) | 2 (11%) | 1 (8%) | 5 (12%) | 1 (8%) |
| **Increased deep tendon reflexes** | 1 (8%) | 1 (6%) | 0 (0%) | 2 (5%) | 0 (0%) |
| **Myoclonus** | 2 (17%) | 2 (11%) | 2 (15%) | 6 (14%) | 0 (0%) |
| **Other neurological diagnoses** |  |  |  |  |  |
| **Acute neurovascular disease** | 0 (0%) | 3 (17%) | 1 (8%) | 4 (9%) | 1 (8%) |
| **Seizures in patients without prior epilepsy** | 0 (0%) | 0 (0%) | 3 (23%) | 3 (7%) | 0 (0%) |
| **Seizures in patients with prior epilepsy** | 2 (17%) | 0 (0%) | 0 (0%) | 2 (5%) | 3 (25%) |

Data are median (IQR), mean (SD) or n/N (%). N is the total number of patients with available data and n the number of patients presenting the finding.

Five patients had overt seizures; two of these had breakthrough seizures attributed to pre-existing epilepsy. Electroencephalogram could not be performed due to restrictions imposed by COVID-19.

Four patients, aged 45 to 59 years, had suffered acute ischemic strokes that could at least partially account for drowsiness. Three of these patients had focal deficits noted in the setting of slow neurologic recovery following IV sedative discontinuation. Of these four patients, two suffered malignant middle cerebral artery infarctions, one had an occipital and midbrain infarct and the other one had a temporal-parietal infarct with hemorrhagic transformation (Table 3).

Laboratory test findings are presented in Table 4, including lymphocytes, platelets, creatinine, blood urea nitrogen (BUN), sodium and C-reactive protein (CRP) for all patients. All encephalopathy patients had CRP above 5 mg/L (Median: 90.2; IQR 49.3 – 174.4 mg/L).

**Table 4.** Laboratory and CSF studies

|  | Mild | Moderate | Severe | Encephalopathy Cases | Cases without Encephalopathy |
| --- | --- | --- | --- | --- | --- |
| **Total** | 12 | 18 | 13 | 43 | 12 |
| **Blood tests values (median)** |  |  |  |  |  |
| **Lymphocytes /mm<sup>3</sup>** | 1,020 (705 – 1,257) | 1,476 (65 – 2,010) | 980 (680 – 1,460) | 1,150 (645 – 1,680) | 1,400 (1,100 – 2,000) |
| **Platelets 10<sup>3</sup>/mm<sup>3</sup>** | 255 (166 – 326) | 270 (213 – 364) | 235 (189 – 254) | 242 (173 – 320) | 292 (218 – 369) |
| **C-reactive protein (mg/L)** | 71.3 (46.7 – 168.3) | 92.7 (45.7 – 156.7) | 115.0 (69.4 – 176.9) | 90.2 (49.3 – 174.4) | 36.4 (11.4 – 176.4) |
| **BUN (mg/dL)** | 22.7 (15.5 – 36.2) | 52.6 (22.4 – 68.1) | 64.5 (28 – 81.8) | 35.5 (21.7 – 81.8) | 13.1 (10.0 – 19.9) |
| **D-dimer (ng/ml)** | 1,974.0 (801.5 – 5,231) | 4,444.0 (1,594 – 14,950) | 4,716.5 (2,411.3 – 14,827.8) | 4,222.0 (1,538.5 – 8,891.3) | 1,469.0 (488.0 – 3,775.3) |
| **Abnormal blood tests** |  |  |  |  |  |
| **Creatinine >1,2(males); >0,9(females) mg/dL** | 5 (42%) | 12 (67%) | 10 (77%) | 27 (63%) | 5 (42%) |
| **Sodium >145mEq/L** | 0 (0%) | 7 (39%) | 3 (23%) | 10 (23%) | 0 (0%) |
| **Sodium <135mEq/L** | 2 (17%) | 2 (11%) | 2 (15%) | 6 (14%) | 1 (8%) |
| **NT-proBNP>125 pg/mL** | 3/3 (100%) | 7/8 (88%) | 4/4 (100%) | 14/15 (93%) | 3/6 (50%) |
| **Creatine phosphokinase >190 (men); >167(women)U/L** | 0/8 (0%) | 6/13 (46%) | 3/11 (27%) | 9/31 (29%) | 3/10 (30%) |
| **ALT > 120 U/L** | 0/12 (0%) | 3/17 (18%) | 0/12 (0%) | 3/41 (7%) | 0/11 (0%) |
| **Indirect bilirubin>0,6 mg/dL** | 0/10 (0%) | 0/16 (0%) | 0/11 (0%) | 0/37 (0%) | 0/10 (0%) |
| **Albumin <3,4 g/dL** | 7/9 (78%) | 13/14 (93%) | 5/7 (71%) | 25/30 (83%) | 6/6 (100%) |
| **Prothrombin time ratio > 1,4** | 0/11 (0%) | 1/16 (6%) | 1/12 (8%) | 2/39 (5%) | 0/10 (0%) |
| **Cerebrospinal fluid findings** |  |  |  |  |  |
| **White blood cells > 4/ mm<sup>3</sup>** | 0/8 (0%) | 1/8 (13%) | 1/7 (14%) | 2/23 (9%) | 0/3 (0%) |
| **Protein > 40 mg/dL** | 2/8 (25%) | 3/8 (38%) | 4/7 (57%) | 9/23 (39%) | 2/3 (67%) |
| **Protein > 40 mg/dL without diabetes** | 0/8 (0%) | 2/8 (25%) | 3/7 (43%) | 5/23 (22%) | 1/3 (33%) |
| **Glucose < 50 mg/dL** | 1/8 (13%) | 1/8 (13%) | 1/7 (14%) | 3/23 (13%) | 0/3 (0%) |
| **Lactate > 22 mg/dL** | 0/8 (0%) | 3/8 (38%) | 2/7 (29%) | 5/23 (22%) | 0/3 (0%) |
| **Elevated CSF Gamma-globulin** | 0/6 | 1/6 (17%) | 0/6 (0%) | 1/18 (5%) | 1/2 (50%) |
| **Oligoclonal bands present** | 0/8 (0%) | 0/8 (0%) | 0/8 (0%) | 0/23 (0%) | 0/3 (0%) |
| **Interleukin 6 > 7,8 pg/mL** | 0/1 (0%) | 1/2 (50%) | 2/2 (100%) | 3/5 (60%) | 0/1 (0%) |
| **PCR for SARS-CoV-2 positive** | 0/6 (0%) | 0/8 (0%) | 1/7 (14%) | 1/21 (4%) | 0/3 (0%) |
| Data are median (IQR), mean (SD) or n/N (%). N is the total number of patients with available data and n the number of positive patients. |  |  |  |  |  |

### Neuroimaging

Thirty-nine of 43 encephalopathy patients underwent neuroimaging studies: brain CT 38, CT angiography 14, brain MRI nine, MRI angiography three. Neuroimaging was normal or showed only chronic changes in 24/38 (63%) of CTs and 4/9 (44%) of MRIs. Twenty-five of the 39 patients (64%) did not present acute changes in neuroimaging studies.

Neuroimaging findings potentially associated with COVID-19 were seen in five patients: acute ischemic stroke in the four described patients and one previously reported case (submitted) displayed multiple rounded T2/FLAIR hyperintensities possibly related to CNS demyelination. Four patients had acute brain imaging abnormalities probably not associated with COVID-19: a pregnant woman with convexity subarachnoid hemorrhage, Wernicke’s encephalopathy findings and pontine myelinolysis, a man with newly diagnosed brain metastasis from lung cancer, a transplant recipient with tacrolimus-associated posterior reversible encephalopathy syndrome, and a patient with a convexity subarachnoid hemorrhage without history of trauma or a related aneurysm.

Five patients with hypertension showed subcortical white matter hypodensities on brain CT; two were confluent, and three, multifocal, none of whom underwent MRI. These findings most likely represented chronic small vessel disease, but the possibility of COVID-19 related changes could not be excluded.

### Cerebrospinal fluid

Lumbar puncture was performed in 23 encephalopathy patients and in three patients without encephalopathy who presented with headache. All three patients without encephalopathy showed normal CSF cell count and mildly elevated protein in two patients (42 and 48 mg/dL).

Among encephalopathy patients, two (9%) had an elevated CSF white blood cell (WBC) count. All 26 samples were negative for oligoclonal bands (Table 4).

Interleukin 6 was quantified in seven CSF samples from six patients (five encephalopathy patients and one non-encephalopathy patient). Interleukin 6 level values were in normal reference ranges (<7.8 pg/mL) in three patients, including one patient without encephalopathy (2.13 pg/mL), one patient with mild encephalopathy (3.44 pg/mL), one with moderate encephalopathy (7.32 pg/mL). Three patients in the encephalopathy group had elevated IL-6 levels. One of these was case report #2 (below), with moderate encephalopathy, and two IL-6 measures (52.99pg/mL and 25.32pg/mL). The two other patients had severe encephalopathy (19.57 – case report #1, below – and 12.15pg/mL).

Case report #1 – SARS-CoV-2 was detected in the CSF of one patient (out of 21): a woman in her 50’s who developed fever during hospitalization for acute aortic dissection complicated by renal failure requiring dialysis, blood eosinophilia, disseminated candidiasis, and central venous catheter-related bloodstream infection. A nasal swab test was positive for SARS-CoV-2. Two days after onset of fever, she developed encephalopathy. A lumbar puncture, obtained ten days after onset of fever, revealed 15 cells/mm3 (38% lymphocytes, 8% monocytes, 22% neutrophils, 31% eosinophils and 1% macrophages), protein of 54 mg/dL, interleukin 6 of 19.57pg/mL (cut-off 7.8 pg/mL) and a positive CSF SARS-COV-RT-PCR. A peripheral blood cell count also revealed elevated leukocytes count with 48% eosinophils. Brain CT was normal, and MRI was not obtained.

Case report #2 – A man over the age of 70 with positive nasal swab SARS-CoV-2-RT-PCR was transferred to our center due to encephalopathy 28 days after respiratory symptom onset.

Brain CT was normal. CSF analysis revealed 256/mm^3^ cells (90% mononuclear cells), a total protein 115 mg/dL, glucose 46 mg/dL, lactate 25.2 mg/dL, and IL-6 52.99 pg/mL. A CSF multiplex PCR assay detected EBV and CSF-SARS-CoV-2-PCR was negative. Additional lumbar punctures were performed three and seven days after the initial exam, and showed persistent CSF pleocytosis, elevated IL-6 (25.32pg/mL) and elevated total protein, but negative EBV-PCR. The patient fully recovered after one month.

### Statistical Analysis

None of the variables showed a correlation with the severity of encephalopathy in the ordinal logistic regression analysis. On secondary analysis, the degree of encephalopathy correlated with higher probability of death during hospitalization; OR of 5.5 for each increment in the degree of encephalopathy (p<0.001).

## Discussion

We report the findings of encephalopathy in 43 patients from 66 neurology consults in 1,263 COVID-19 patients. The presentation of encephalopathy ranged from behavioral changes and mild attention deficit to severely impaired consciousness. Its degree correlated with mortality

Our series represents cases in the COVID-19 dedicated center presenting with more severe neurological manifestations, or those in which clinicians could not attribute symptoms to systemic dysfunctions alone. These cases would represent the sample of patients most likely to present neurological disease caused by direct central nervous system viral invasion. However, evidence of neuroinvasion and neuroinflammation were largely absent. Although clinical factors, including renal impairment and prolonged sedative use were highly prevalent in moderate and severe encephalopathy cases, many patients displayed encephalopathy without clear metabolic dysfunction or sedative use, especially in mild encephalopathy cases. The only consistent findings in these patients were laboratory signs of systemic inflammation.

Neurological examination showed few additional findings other than encephalopathy. Some patients presented myoclonus, attributed to toxic and metabolic disorders. In two patients abnormal movements noted on neurological exam suggested focal seizures. Only one patient in this series presented severe psychomotor agitation on examination. In other patients, agitation had been controlled with antipsychotics and intravenous sedatives by the time of the neurology evaluation.

The moderate encephalopathy group was the largest group in our series, comprising patients seen later in the disease course, many of whom had spent prolonged periods under sedation and mechanical ventilation. These patients were easily arousable, but had severely compromised level of attention, in some cases unable to engage in basic attention testing.

Although encephalopathy is a common finding in COVID-19 inpatients, the precise underlying mechanisms remain incompletely understood. Case reports or small case series that evaluated SARS-CoV-2 RNA detection in CSF samples showed conflicting results: while some studies detected viral RNA with RT-PCR or next-generation sequencing ^10-14^, other studies reported negative findings ^15-20^. Generalizability of findings is possibly limited by publication bias. Our study included 24 CSF SARS-CoV-2-RT-PCR tests. We found only one SARS-CoV-2-positive sample, which may have resulted from shedding of viral genetic material from plasma to CSF in a patient with damaged blood-brain barrier. Our findings suggest that SARS-CoV-2 RNA detection rates in encephalopathy cases may be lower than expected from case reports, but consistent with other larger case series^6^. Other studies demonstrated the presence of anti-SARS-CoV-2 antibodies in the CSF of patients with neurologic disease and negative CSF-RT-PCR ^19,20^, suggesting that immunological assays may be superior to molecular testing to demonstrate SARS-CoV-2 CNS involvement. In other infectious diseases, such as Lyme neuroborreliosis, neurosyphilis, West Nile virus encephalitis and Japanese encephalitis, immunological assays appear to be superior to molecular testing in blood or CSF, which may be related to low pathogen loads or transient CNS infections ^21^.

Alternatively, encephalopathy may be caused by CNS immune-mediated processes without viral invasion. Our series is in agreement with larger studies showing that CSF cell and biochemistry counts are largely normal in COVID-19 patients with neurological symptoms ^5,6,19^ However, elevated CSF WBC, protein levels, and positive oligoclonal bands were described in case reports^3^. Oligoclonal bands were absent in all 24 CSF samples in our series. Only two patients (8.3% of cases) showed increased CSF WBC. In both cases alternative explanations not related to SARS-CoV-2 CNS infection (case reports #1 and #2) were postulated.

Cytokine storm is a potential major cause of acute respiratory distress syndrome and multiple organ dysfunction in COVID-19 patients ^22^. Cytokine serum levels, including IL-1, IL-6 and TNF-α, correlate with systemic COVID-19 severity ^23,24^ Recent studies demonstrated elevated CSF IL-6 levels in patients with encephalopathy, negative CSF SARS-CoV-2-RT-PCR and normal CSF cell count, suggesting a potential role of IL-6 as a biomarker in these patients ^20,25^. We found markedly elevated IL-6 levels in two patients with known inflammatory CNS disease, both of which were possibly not COVID-related (case reports #1 and 2#).

Although most COVID-19 neuroimaging studies evaluated patients with acute cerebrovascular disease, suspected myelopathy and miscellaneous neurologic syndromes, neuroimaging was requested to evaluate mental status changes in 37-73% of patients ^26-29^. In our series, the majority of patients had brain CTs or MRIs, most of which did not display acute changes. Acute infarcts (10.3% of neuroimaging studies) and white matter abnormalities suggestive of microangiopathy (12.8%) were the most common findings, in keeping with other retrospective studies ^28,29^. Only one case, reported elsewhere (submitted), had acute demyelinating-like lesions potentially related to COVID-19. In other COVID-19 patients neuroimaging findings could be explained by other neurological diseases, however their association with COVID-19 cannot be completely ruled out. In our series we did not observe other previously described neuroimaging findings, such as punctate hemorrhagic foci, cortical swelling and leptomeningeal enhancement^30^. Since only a few patients in this series underwent MRI, these findings could be missed on brain CT.

We found subcortical white matter abnormalities on brain CT in five Covid-19 patients, all with a prior history of hypertension. It remains unclear if these changes are related to previous hypertensive small vessel disease or if they could be related to COVID-19. These abnormalities could be particularly common in COVID-19, and not easily distinguishable from hypoxic leukoencephalopathy and other toxic-metabolic causes ^27,31^.

## Conclusion

In our series, encephalopathy in COVID-19 manifested in various degrees of severity which were associated with increased mortality and may be influenced by factors such as sedative drug use, organ system dysfunction and systemic inflammation. Head CT showed mainly non-specific changes, and MRI may be preferable, though not easily obtainable in critically ill patients, to document more subtle findings related to encephalopathy in COVID-19 patients. CSF analysis in our series did not detect viral genetic material or inflammatory changes, suggestive of CNS viral invasion or immune-mediated processes, in the majority of encephalopathy cases. Future studies should clarify the role of cytokines, CSF-SARS-CoV-2 immunology and other inflammatory biomarkers, particularly in mild and moderate cases of COVID-19 associated encephalopathy.

## Data Availability

I take full responsibility for the data, the analyses and interpretation, and the conduct of the research. I have full access to all of the data and have the right to publish any and all data separate and apart from any sponsor.

## Declarations

Dr. Tuma, Dr. Guedes, Dr. Carra, Dr. Iepsen, Dr. Rodrigues, Dr. Camelo-Filho, Dr. Kubota, Dr. Ferrari, Dr. Studart Neto, Dr. Oku, Dr. Terrim, Dr. Lopes, Dr. Passos Neto, Dr. Fiorentino, Dr. Souza, Dr. Baima, Dr. Silva, Dr. Perissinotti, Dr. Martin, Dr. Gonçalves, Dr. Fortini, Dr. Smid, Dr. Adoni, Dr. Lucato, Dr. Nitrini, Dr. Gomes and Dr. Castro report no disclosures.

This study has not been sponsored.

The corresponding author takes full responsibility for the data, the analyses and interpretation, and the conduct of the research. He has full access to all of the data and has the right to publish any and all data separate and apart from any sponsor.

As stated in the Methods section of the manuscript, this study was approved by the hospital’s ethics committee.

There is no conflict of interests to declare.

